# Deep learning enables fully automated cineCT-based assessment of regional right ventricular function

**DOI:** 10.1101/2025.09.29.25336897

**Authors:** Amanda Craine, Kaiden Simon, Lauren Severance, Laith Alshawabkeh, Nick H. Kim, Eric Adler, Anna Narezkina, Ori Ben-Yehuda, Francisco Contijoch

## Abstract

**Background:** Right ventricular (RV) function is a key factor in the diagnosis and prognosis of heart disease. However, current advanced CT-based assessments rely on semi-automated segmentation of the RV blood pool and manual delineation of the RV free and septal wall boundaries. Both of these steps are time-consuming and prone to inter- and intra-observer variability.

**Methods:** We developed and evaluated a fully automated pipeline consisting of two deep learning methods to automate volumetric and regional strain analysis of the RV from contrast-enhanced, ECG-gated cineCT images. The Right Heart Blood Segmenter (RHBS) is a 3D high resolution configuration of nnU-Net to define the endocardial boundary, while the Right Ventricular Wall Labeler (RVWL) is a 3D point cloud-based deep learning method to label the free and septal walls. We trained our models using a diverse cohort of patients with different RV phenotypes and tested in an independent cohort of patients with aortic stenosis undergoing TAVR.

**Results:** Our approach demonstrated high accuracy in both cross-validation and independent validation cohorts. RHBS and RVWL both yielded Dice scores of 0.96, and accurate volumetry metrics. RVWL achieved high Dice scores (>0.90) and high accuracy (>93%) for wall labeling. The combination of RHBS and RVWL provided accurate assessment of free and septal wall regional strain, with a median cosine similarity value of 0.97 in the independent cohort.

**Conclusions:** A fully automated 3D cineCT-based RV regional strain analysis pipeline has the potential to significantly enhance the efficiency and reproducibility of RV function assessment, enabling the evaluation of large cohorts and multi-center studies.

**Key Points:**

1. RV endocardial segmentation of contrast enhanced CT scans can be utilized to perform volumetry, and when paired with labeling of free and septal walls, regional evaluation of surface strain.
2. However, this has previously been performed using time-intensive semi-automated segmentation methods and manually labeling free wall and septal wall regions..
3. Here, we describe an automated, deep learning-based approach which uses two separate DL models to define the endocardial boundary (in 3D) and then label the free and septal walls on the endocardial surface.
4. Our approach facilitates rapid and automatic advanced phenotyping of patients. This reduces prior limitations of potential interobserver variability and challenges associated with evaluating large cohorts.

## Introduction

Right ventricular (RV) volumetry and systolic performance is an essential metric for diagnosis and prognosis of both right- and left-sided heart disease. For example, RV volumetry is an important indication for intervention of valvular disease (1–3), and long-term management of congenital heart disease like tetralogy of Fallot (4,5). Further, pre-operative RV dysfunction in the presence of left ventricular (LV) failure has been shown to be predictive of RV failure after LVAD implantation (6,7). In addition to volumetry-based metrics, our group has used ECG-gated cineCT to estimate metrics of regional RV strain and evaluate patients with various pathophysiologies of RV dysfunction (8–10). These detailed assessments highlight unique features of patient-specific preload and afterload that contribute to RV function.

However, current assessments rely on semi-automated segmentation of the RV bloodpool for delineation of the endocardial boundary and manual delineation of the RV free and septal wall boundaries (8–10). The lack of automated analysis limits clinical translation; it leads to inter- and intra-observer variability, limits the scale of datasets that can be evaluated, and hinders multi-center study. Several technical factors impair the routine segmentation of the RV blood pool and labeling of free and septal walls including the complex, crescent-like shape of the RV, the high prevalence of metal artifacts due to replacement valves or pacing wires, and the need for high spatial resolution segmentation for downstream regional evaluation.

Recently, deep learning algorithms have enabled automated 3D cardiac image segmentation with high accuracy and reproducibility. However, a method for RV segmentation for regional processing has not yet been reported. Current algorithms have either focused on left heart structures (11,12), did not include a wide range of disease phenotypes (13–15), or analyzed cardiac MR images (16,17). Further, methods which did delineate the RV endocardial boundary often performed 2D slice-by-slice segmentation, predicted smooth boundaries from 3D volumes or significantly downsampled the imaging volume (13,16,17). However, nnU-Net is a robust semantic segmentation approach that adapts to a given dataset and generates a U-Net segmentation model based on the dataset provided (18). Here, we utilize nnU-Net V2 (19) to delineate the RV endocardial boundary from a heterogeneous dataset of patients who underwent ECG-gated cineCT. We evaluate the accuracy of our method, Right Heart Blood Segmenter (RHBS), using volumetric measures as well as the ability to provide comparable strain metrics (9,20,21).

To interpret regional strain differences within the RV, delineation of the free and septal walls is needed (9,10). Previously, surfaces-of-interest were manually delineated at the end-diastolic timeframe. Deep learning architectures have been developed which can label 3D point cloud data (22,23). However, to date, such an approach has not been used to delineate the RV free and septal walls. Here, we present such an approach - Right Ventricular Wall Labeler (RVWL) - and evaluate the accuracy of the automated labeling as well as its impact on downstream strain analysis.

In this work, we develop and evaluate a fully automated pipeline which consists of two deep learning methods to automate volumetric and regional strain analysis of the RV from contrast enhanced, ECG-gated cineCT images. We address three unanswered questions: 1) Can a 3D convolutional neural network, RHBS, trained on a heterogeneous set of patients yield accurate estimates of high resolution, volumetric segmentations of the RV bloodpool from which volume and regional strain can be accurately measured? 2) Can a 3D point cloud-based deep learning method, RVWL, accurately label the RV free and septal walls of the bloodpool segmentation? 3) Does the combination of the two methods RHBS+RVWL provide accurate assessment of free and septal wall regional strain? After cross-fold validation, we tested our methods in an independent dataset of patients scanned as part of the work-up for transcatheter aortic valve replacement at our institution. This new population was used to evaluate the extent to which the deep learning methods provided accurate automated analysis.

## Methods

The overall data processing pipeline is shown in **Figure 1**. The utility of such an automated pipeline is shown in **Figure 2**.

**Figure 1:**
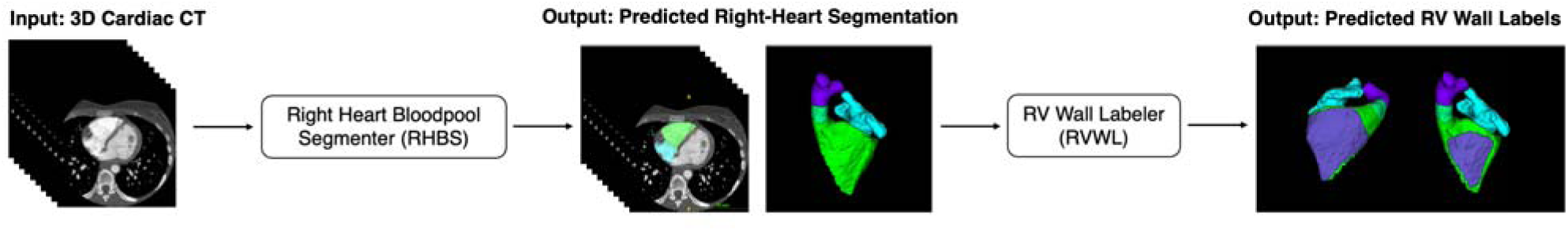
Overview of processing pipeline. Right Heart Bloodpool Segmenter (RHBS) is a nnU-Net V2 based DL model trained to automatically segment the RV, RA, pulmonic valve (PV), and PA bloodpools from ECG-gated cineCT images. Then, RV Wall Labeler (RVWL) delineates the free and septal walls of the RV bloodpool boundary.

**Figure 2:**
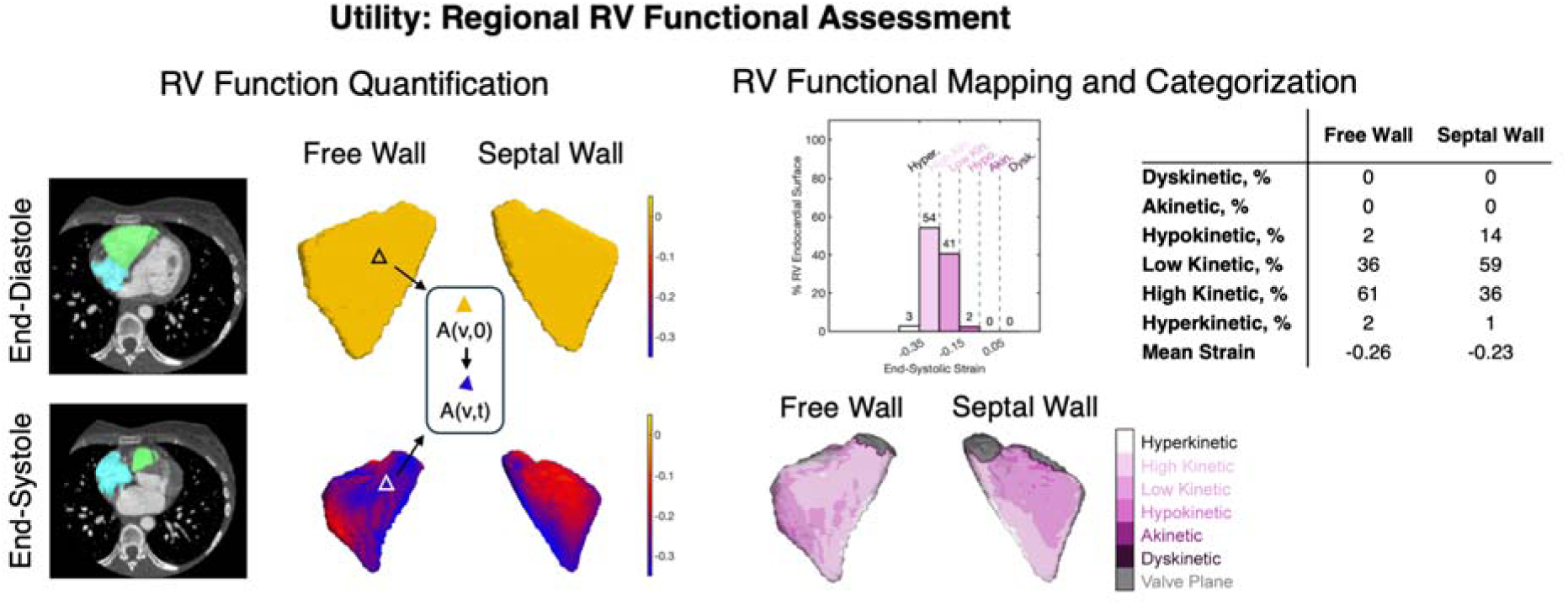
Example of Quantitative Evaluation of RV Free and Septal Wall Strain. Endocardial surface tracking from RV segmentation yields a 3D map of regional strain (left). Each patch on the surface is then categorized into one of 6 groups based on the end-systolic strain value to obtain a profile of strain per patient (histogram and map below). Strain profiles for the free wall and septal wall are obtained by reporting the strains from the patches included in the predicted free wall and septal wall labels (table on right).

### Training Population

With IRB approval, 101 ECG-gated, contrast-enhanced cardiac CT scans obtained between 07/2017 and 11/2023 which were manually processed as part of earlier studies were used for model training (6,7,9,10). Cases used in training were identified from several clinical populations who routinely undergo cardiac cineCT at our institution: patients undergoing anthracycline therapy (AN, n = 8), pre-operative assessment of chronic thromboembolic pulmonary hypertension (CTEPH, n = 14), left-sided heart failure patients undergoing evaluation for LVAD implantation (LVHF, n = 50), and adults with congenital heart disease undergoing evaluation for transcatheter pulmonary valve replacement (ACHD, n = 29).

All patients underwent full cycle, ECG-gated cineCT on a 256-slice Revolution CT scanner (Waukesha, WI). Gantry rotation speed was 280 ms for nearly all cases (n = 1 acquired at 234 ms). Based on the clinical imaging protocol, patients were scanned at 80, 100, or 120 kV and the maximum tube current ranged from 137 - 781 mA. Nearly all images were reconstructed with 0.625 mm slice thickness (n = 6 reconstructed with 1.25 mm slices). Most axial images were reconstructed at ∼10% intervals (n = 23 reconstructed at <9% intervals) of the cardiac cycle (0 - 90% of the R-R interval). However 6 cases covered 0 - <85% of the cardiac cycle. Images were reconstructed on a 512x512 matrix using a standard reconstruction kernel. The field of view was typically 200 mm (range: 179 - 360 mm), which led to an in-plane pixel size of 0.39x0.39mm. RV enhancement ranged from 203 - 1057 Hounsfield units. All cases had sufficient image quality for manual segmentation. Images were not excluded due to the presence of metal artifact (n=49 had metal artifacts present in the RV).

Two frames per patient were selected for training; the end-diastolic frame and an estimation of the end-systolic frame based on the total number of frames. Therefore, the total training dataset consisted of 202 images.

### Test population

Model accuracy was also evaluated in an independent cohort of patients with severe aortic stenosis undergoing evaluation for transcatheter aortic valve replacement (n = 24). Scans occurred between 12/2019 - 11/2022. After review of all scans (n = 290), 24 cases were selected to achieve a balanced distribution of cases with varying RV enhancement.

All patients underwent full cycle, ECG-gated cineCT on a 256-slice Revolution CT scanner (Waukesha, WI). Gantry rotation speed was 280 ms. Based on the clinical imaging protocol, patients were scanned at 80, 100, or 120 kV and the maximum tube current ranged from 402 - 719 mA. Nearly all images were reconstructed with 0.625 mm slice thickness (n = 1 reconstructed with 1.25 mm slices). 11 cases had axial images reconstructed at ∼10% intervals of the cardiac cycle, while 13 cases were reconstructed at <9% intervals. Images were reconstructed on a 512x512 matrix using a standard reconstruction kernel. The field of view was typically 250 mm (range: 190 - 271 mm), which led to an in-plane pixel size of 0.49x0.49mm. RV enhancement ranged from 218 - 1207 Hounsfield units. Seven cases had metal artifacts present in the RV.

### Semi-automated expert bloodpool segmentation and metrics of RV function

Voxel-wise manual segmentation of the RV, right atrium (RA), pulmonic valve (PV), and pulmonary artery (PA) blood volumes on images at the native resolution were confirmed by a trained operator with 6 years of experience in cardiac image segmentation (author A.C.) using ITK-SNAP (Philadelphia, PA). RV volumes were obtained from each segmentation. Global RV function was measured as end-diastolic volume (EDV) and end-systolic volume (ESV), while regional RV function was measured using regional strain (RS_CT_). Mean RV RS_CT_ and mean RS_CT_ in the free and septal walls were measured. RS_CT_ values were also categorized by the distribution of strain strength at end-systole. Methods for measuring and categorizing RV RS_CT_ from cardiac cineCT have been previously described in detail (8,9).

### Automated Delineation of the RV Endocardial Volume/Boundary via RHBS

For uniformity, images were resampled to 1 mm isotropic voxels. RHBS labeled each voxel as one of five labels: RV, RA, PV, PA, and background. We trained the model using nnU-Net V2’s 3D full resolution configuration (19) on a 16-core Ubuntu computer with 128 GB RAM and a 24 GB NVIDIA RTX A5000 (NVIDIA Corporation, Santa Clara, CA). Training and validation were performed using five-fold stratified cross-validation to ensure each fold included a representative sample of each clinical population. Each fold was trained for 1000 epochs per nnU-Net default.

### Metrics of RHBS Accuracy

RV segmentation accuracy was evaluated volumetrically with the Dice coefficient. Global volumetry metrics were evaluated using the absolute and percent errors of RV end-diastolic and end-systolic volumes as well as the difference in mean end-systolic RV strain measured with the predicted segmentations relative to the manual approach.

Regional function accuracy was measured as the cosine similarity between the end-systolic RS_CT_ categorized with the manual and predicted segmentations. Cosine similarity measures the similarity between the directions of two vectors in space. Given that the RS_CT_ categorization is reported as the percent of the RV surface that belongs to each category (9), this distribution was treated as a vector in multi-dimensional space. Cosine similarity evaluated the similarity of the direction of two vectors, in this case the vectors described the RS_CT_ categorizations of the reference segmentation and RHBS predicted segmentation.

### Manual Labeling of Free and Septal Walls

The end-diastolic phase of the ECG-gated cineCT dataset was used for labeling of the free and septal wall surfaces (9,10). To do so, the semi-automated segmentation of the RV blood pool was visualized using ITK-SNAP (Philadelphia, PA). Each surface was then delineated by a trained operator with 6 years of experience in cardiac image segmentation (author A.C.). The labeling does not affect volumetric or global strain analysis. It is used to select the portions of the RV endocardial mesh for analysis of regional strain.

### Automated Labeling of Free and Septal Walls via RVWL

The semi-automatic RV bloodpool segmentations with free wall and septal wall labels were converted into point clouds for automated free and septal wall labeling. Each patient yielded a set of surface points that were labeled as either being the “free wall”, “septal wall”, or “none”.

PointNet’s architecture maps each independent point within a point cloud to a high-dimensional feature and uses maxpooling to aggregate these features into a global descriptor (22). PointNet++ extends this approach by introducing hierarchical set abstraction which enables the model to learn local geometric features with spatial coherence (23). Multi-Scaling Grouping (MSG) creates several neighborhoods that each have a different radius, allowing PointNet++ to capture more detailed information about both fine-grained local structures and broader contextual geometry of the Free Wall and Septal Wall.

RVWL is a PointNet++ with MSG network trained to predict the labels generated by human annotation. Each scan yields 1 training example so the total training dataset was 101 point clouds. PointNet++ analyzes 3,000 points (from a point cloud) at a time but our surfaces had between 11,405 and 36,789 points. Therefore, surface points were subsampled into 3,000 point sets which served as an augmentation during training. The network was trained using the PyTorch for 1000 epochs. The same cross-fold partitions defined for the 3D U-net were used for PointNet++ training and cross-fold evaluation.

Post-processing of predictions was performed to minimize mislabeling of the walls due to weak predictions. Specifically, after prediction, free wall and septal wall labels were only kept if the class label probability ≥ 0.9. Otherwise, these weak predictions were relabeled as “none”.

Wall labeling performance was evaluated with the Dice score of each label (free wall, septal wall, none). Additionally, overall accuracy and extent of mislabeling (i.e. a true septal wall point predicted as free wall) was also evaluated. Accuracy of resulting regional function was measured as the correlation of mean end-systolic strain of the free wall and septal wall with respect to the standard approach. Regional function accuracy was also measured as the cosine similarity between the predicted strain profiles for each wall and the standard approach.

### Statistics

Normality of datasets was evaluated with the Shapiro-Wilk test. For nonparametric datasets, statistical differences in prediction results between the cross validation cohort and the independent cohort were evaluated with a two-sided Wilcoxon rank sum test. Correlation studies between nonparametric datasets were computed with the Spearman correlation. Pearson correlation was used to evaluate normally distributed data. Statistical testing was computed with MATLAB (MathWorks, Natick, MA).

## 3) Results

### Patient demographics

Patient demographics are summarized in **Table 1**. TAVR recipients were older (83 years, IQR: 74 - 91) than the AN (59 years, IQR: 40 - 69, p = 0.01), CTEPH (57 years, IQR: 49 - 61, p < 0.01), LVHF (59 years, IQR: 50 - 68, p < 0.01), and ACHD (32 years, IQR: 24 - 35, p < 0.01) cohorts. The ACHD patients were younger than the AN (p = 0.04), CTEPH (p < 0.01), and LVHF (p < 0.01) patients.

**Table 1:** Demographic and distribution of patients amongst training folds. TAVR recipients were older than the AN, CTEPH, LVHF, and ACHD cohorts while the ACHD cohort was younger than the AN, CTEPH, and LVHF cohorts. The LVHF cohort had fewer women than the AN, CTEPH, and ACHD cohorts, and had higher heart rates than the AN and TAVR cohorts. Five-fold cross validation distributed the populations across the five folds as evenly as possible.

|  | Cross-Fold Training and Validation |  |  |  | Indep. Validation |  |  |
| --- | --- | --- | --- | --- | --- | --- | --- |
|  | AN<br>N = 8 | CTEPH<br>N = 14 | LVHF<br>N = 50 | ACHD<br>N = 29 | TAVR<br>N = 24 | Total<br>N = 125 | p-value |
| **Demographics** |  |  |  |  |  |  |  |
| **Age, years** | 59<br>(40 - 69) | 57<br>(49 - 61) | 59<br>(50 - 68) | 32<br>(24 - 35) | 83<br>(74 - 91) | 57<br>(40 - 70) | **<0.01** |
| **Sex, n (%)** | 7 (88) | 8 (57) | 6 (12) | 14 (48) | 9 (38) | 44 (35) | **<0.01** |
| **HR (bpm)** | 61<br>(53 - 66) | 76<br>(65 - 80) | 81<br>(70 - 95) | 72<br>(61 - 81) | 63<br>(57 - 76) | 74<br>(63 - 86) | **<0.01** |
| **Distribution Amongst Folds** |  |  |  |  |  |  |  |
| **Fold 1, n (%)** | 2 (10) | 3 (14) | 10 (50) | 6 (29) | - | 21 (21) | - |
| **Fold 2, n (%)** | 2 (10) | 3 (15) | 10 (50) | 5 (25) | - | 20 (20) | - |
| **Fold 3, n (%)** | 2 (10) | 2 (10) | 10 (50) | 6 (30) | - | 20 (20) | - |
| **Fold 4, n (%)** | 1 (5) | 3 (15) | 10 (50) | 6 (30) | - | 20 (20) | - |
| **Fold 5, n (%)** | 1 (5) | 3 (15) | 10 (50) | 6 (30) | - | 20 (20) | - |

The LVHF cohort had fewer women (n = 6/50) than the AN (n = 7/8, p < 0.01), CTEPH (n = 8/14, p = 0.02), and ACHD (n = 14/29, p = 0.01) cohorts.

Median heart rate was 74 beats per minute (IQR: 63 - 86). The LVHF cohort had higher heart rates (81 bpm, IQR: 70 - 95) than the TAVR (63 bpm, IQR: 57 - 76, p < 0.01) and AN (61 bpm, IQR: 53 – 66, p = 0.03) cohorts.

### RHBS training and quantitative evaluation

#### Training

Folds 1 was trained on 80 cases and validated on 21 cases (160 and 42 volumes, respectively). Folds 2 - 5 were trained on 81 cases and validated on 20 cases (162 and 40 volumes, respectively). The distribution of each population within each fold is demonstrated in Table 1. The independent validation dataset was predicted using the model from Fold 3.

#### Quantitative Evaluation

RHBS performance results are shown in **Table 2**. Dice was high and comparable in the cross validation (0.96, IQR: 0.95 - 0.97) and independent validation cohorts (0.96, IQR: 0.94 - 0.97, p > 0.05). Absolute volume error and percent volume error were not different between the cross validation (absolute error: 7 mL, IQR: 3 - 12; percent error: 4%, IQR: 2 - 8) and independent validation cohorts (absolute error: 5 mL, IQR: 2 - 9, p > 0.05; percent error: 5%, IQR: 3 - 7, p > 0.05). 95% of cases were segmented with Dice > 0.90 and percent volume error < 20%. End-systolic frames had lower Dice scores than end-diastolic frames for both the cross validation (end-diastole: 0.97, IQR: 0.96 - 0.98; end-systole: 0.95, IQR: 0.94 - 0.96; p < 0.01) and independent validation (end-diastole: 0.97, IQR: 0.96 - 0.97; end-systole: 0.94, IQR: 0.93 - 0.96; p <0.01) cohorts (**Figure 3**). We also evaluated the impact of RV enhancement of RHBS accuracy and did not find a significant effect (**Figure 3**).

**Figure 3:**
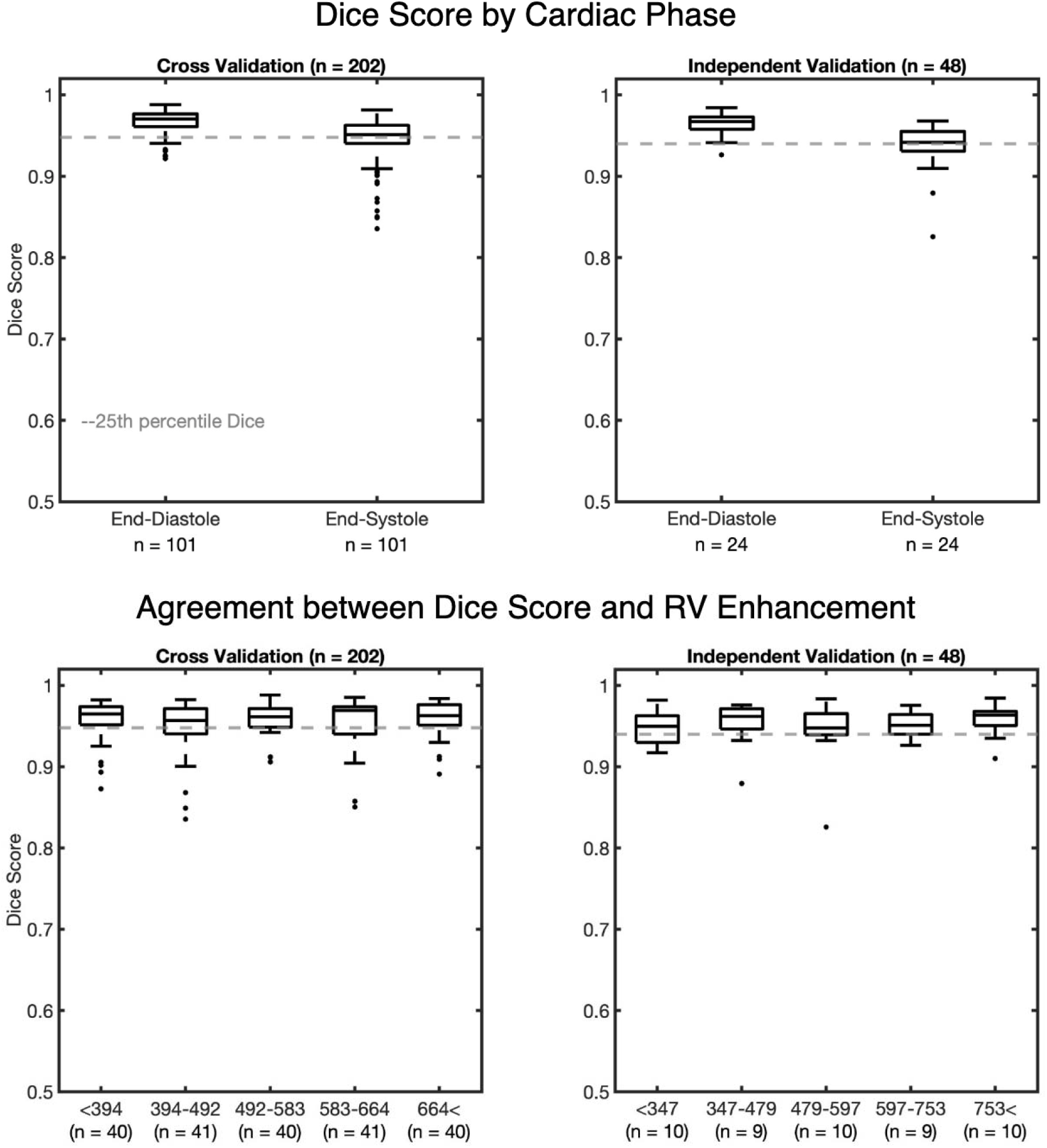
Dice score evaluated by cardiac phase and level of RV enhancement. End-diastolic frames had higher Dice scores than the end-systolic frames in both the cross validation and independent validation cohorts. Dice scores were not different across quintiles of RV enhancement in the cross validation or independent validation cohorts. Gray dashed line indicates the 25th percentile of Dice scores for each dataset.

**Table 2:** Right Heart Blood Segmenter (RHBS) cross-validation and Independent Validation Result Metrics. Dice scores, absolute volume error, percent volume error, and cosine similarity were similar in the cross validation and independent validation cohorts.

| Metric | Cross Validation<br>(n = 202) | Independent Validation<br>(n = 48) | p-value |
| --- | --- | --- | --- |
| Dice Score | 0.96 (0.95 - 0.97) | 0.96 (0.94 - 0.97) | 0.06 |
| Absolute Volume Error (mL) | 7 (3 - 12) | 5 (2 - 9) | 0.07 |
| % Volume error | 4 (2 - 8) | 5 (3 - 7) | 0.08 |
| Cosine similarity | 0.98 (0.95 - 0.99) | 0.99 (0.95 - 0.99) | 0.10 |

RHBS yielded mean RV strain measures which agreed with values derived from semi-automated, expert segmentations (**Figure 4, left**). Pearson correlation was 0.97 for the cross validation cohort and 0.98 for the independent validation cohort, and were not significantly different from one another (p = 0.54). The root mean squared error was 0.02 and 0.01 for the cross validation and independent validation cohorts respectively.

**Figure 4:**
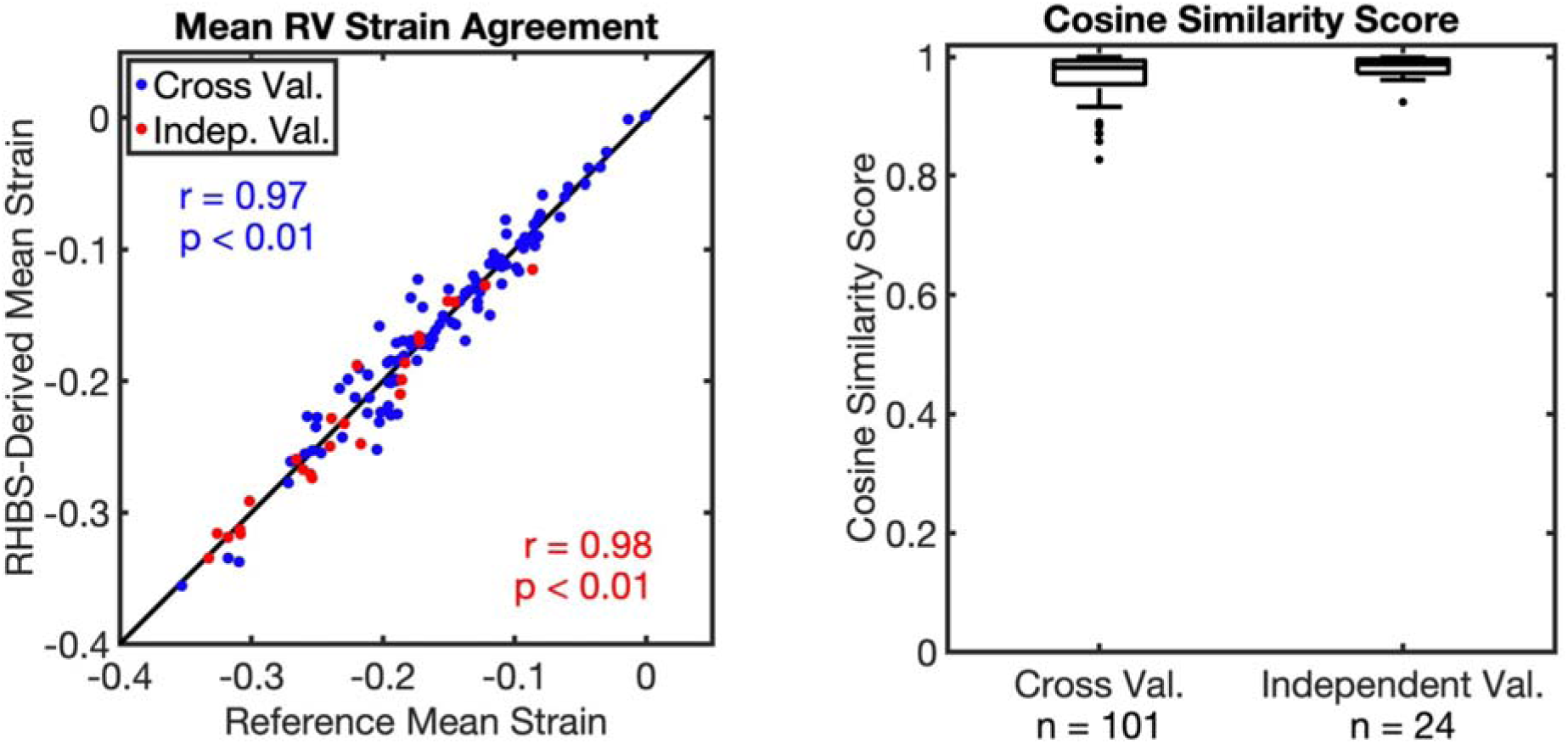
Accuracy of DL-based Right Heart Bloodpool Segmenter (RHBS)-derived mean RV strain (left) and strain categorization (right). **Left:** DL-derived segmentation agreed very strongly in both the cross-validation (blue) and independent validation (red) cohorts with semi-automated RV segmentation based analysis. **Right:** In addition to overall mean strain, categorization of the RV strain very closely (>0.98) agreed with semi-automated analysis and was comparable between both validation cohorts.

RHBS yielded accurate categorization of RS_CT_. Cosine similarity (**Figure 4, right**) was similar between both testing cohorts (cross-fold validation: 0.98, IQR: 0.95 - 0.99; independent: 0.99, IQR: 0.97 - 0.99, p > 0.05).

### RVWL training and quantitative evaluation

#### Training

The best performing RVWL for each fold had a validation dice of 0.91 or 0.92, and occurred between epoch 322 and 889. The independent validation dataset was predicted using the model from fold 3.

#### Quantitative Evaluation

**Table 3** summarizes quantitative evaluation of RVWL. Dice and mesh accuracy for all labels across both cross-validation and independent testing cohorts was high (>0.90 and >93%, respectively). Mislabeling (SW points labeled FW and vice-versa) on the RV mesh was low (≤1%), however, independent validation had a small, but statistically greater number of septal wall points mislabeled as free wall (1%, IQR: <1 - 2) compared to the cross-validation dataset (<1%, IQR: 0 - 1, p<0.01). RVWL applied to manual segmentations resulted in high CSM (0.99 in both the cross-fold and independent testing cohorts).

**Table 3:** Right Ventricle Wall Labeler (RVWL) Cross-Validation and Independent Validation Result Metrics. Dice and accuracy of point labeling was high across walls and validation groups. Percentage of points mislabeled (either true septal wall point labeled free wall or vice-versa) was low although statistically higher in the independent validation for SW points. Cosine similarity metric (CSM) of strain analysis was high across walls and cohorts for both RVWL based labeling and the combination of RHBS segmentation followed by RVWL labeling.

| Metric |  | Cross validation<br>(n = 101) | Independent validation<br>(n = 24) | p-value |
| --- | --- | --- | --- | --- |
| RVWL Dice | FW | 0.94 (0.92 - 0.95) | 0.93 (0.91 - 0.94) | 0.13 |
|  | SW | 0.92 (0.90 - 0.94) | 0.93 (0.92 - 0.94) | 0.68 |
|  | None | 0.90 (0.88 - 0.92) | 0.91 (0.89 - 0.92) | 0.67 |
| RVWL Mesh Accuracy | FW | 95 (93 - 97) | 96 (93 - 98) | 0.31 |
|  | SW | 94 (91 - 97) | 93 (89 - 95) | 0.06 |
| RVWL % Mislabeled | FW | 0 (0 - <1) | 0 (0 - <1) | 0.48 |
|  | SW | <1 (0 - 1) | 1 (<1 - 2) | **<0.01** |
| RVWL CSM | FW | 0.99 (0.99 - 0.99) | 0.99 (0.99 - 0.99) | 0.12 |
|  | SW | 0.99 (0.99 - 0.99) | 0.99 (0.99 - 0.99) | 0.06 |
| RHBS+RVWL CSM | FW | 0.97 (0.93 - 0.99) | 0.97 (0.94 - 0.99) | 0.77 |
|  | SW | 0.97 (0.94 - 0.99) | 0.97 (0.93 - 0.98) | 0.20 |

Subsequently, the combination of RHBS and RVWL was evaluated. Mean RV strain derived from RHBS+RVWL was very strongly correlated to mean strain derived from the semi-automated approach (**Figure 5, top**). Mean strain Pearson correlations were not different between the cross validation or independent validation cohorts in the free wall (Cross: r = 0.96, p < 0.01; Indep: r = 0.95, p < 0.01; r-to-z transformation p = 0.85) or septal wall (Cross: r = 0.96, p < 0.01; Indep: r = 0.93, p < 0.01; r-to-z transformation p = 0.78). CSM for the combined approach of RHBS+RVWL is reported in **Table 3**. Median CSM was 0.97 in the independent validation. The accuracy of the full automated approach is further illustrated in **Figure 5, bottom.**

**Figure 5:**
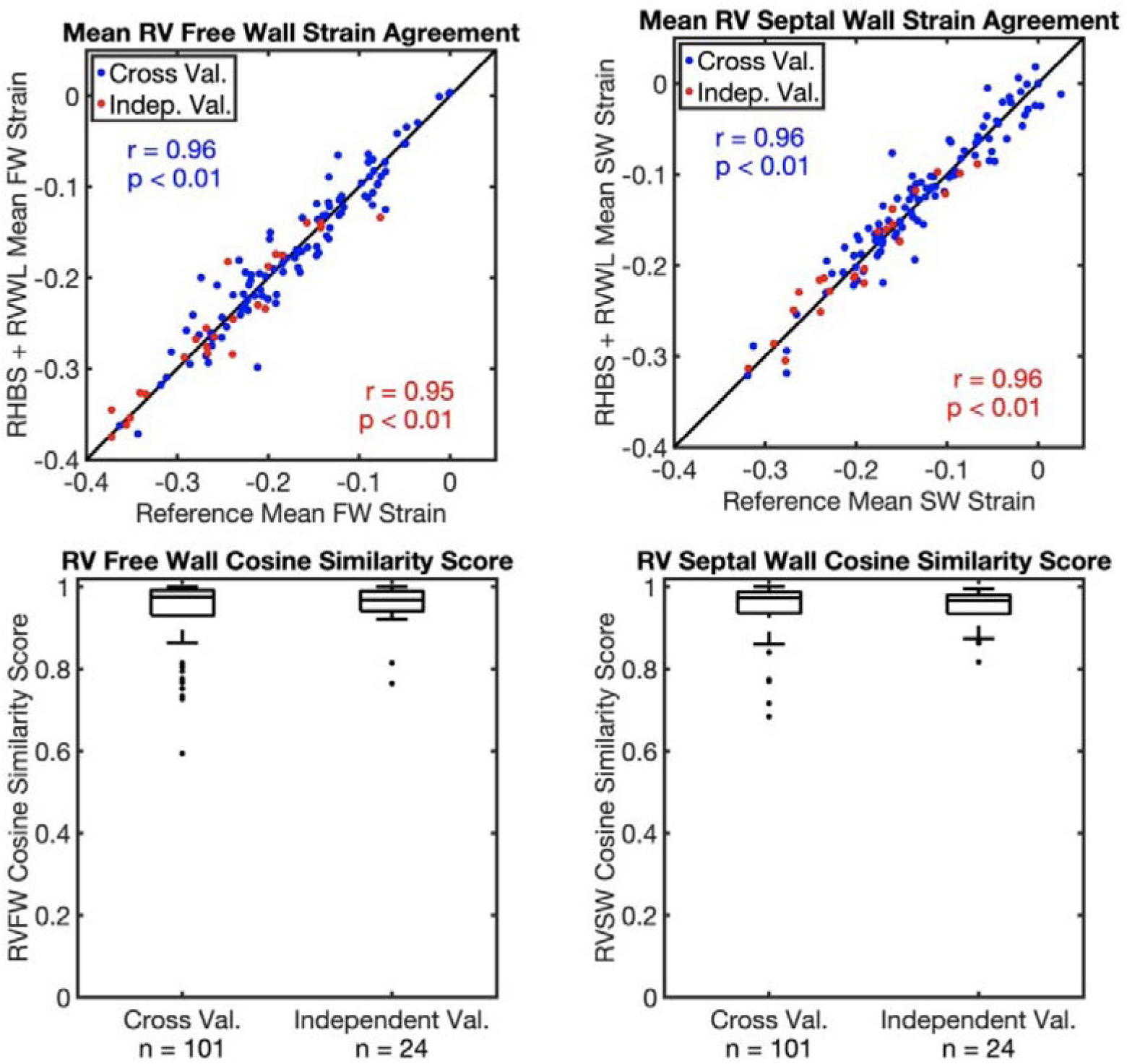
Accuracy of RHBS+RVWL-derived FW and SW strain in both cross-fold validation and independent testing cohort. **Top:** The combination of both DL-based RHBS and DL-based RVWL provides mean FW and SW strain estimates which closely match standard semi-automated processing. **Bottom:** Classification of surface strain into different categories with RHBS+RVWL closely matches the reference method.

#### Representative Examples

Automated RV volumetry and strain mapping for two representative patients is shown in **Figure 6** (left: cross validation example, right: independent validation cohort example).

**Figure 6:**
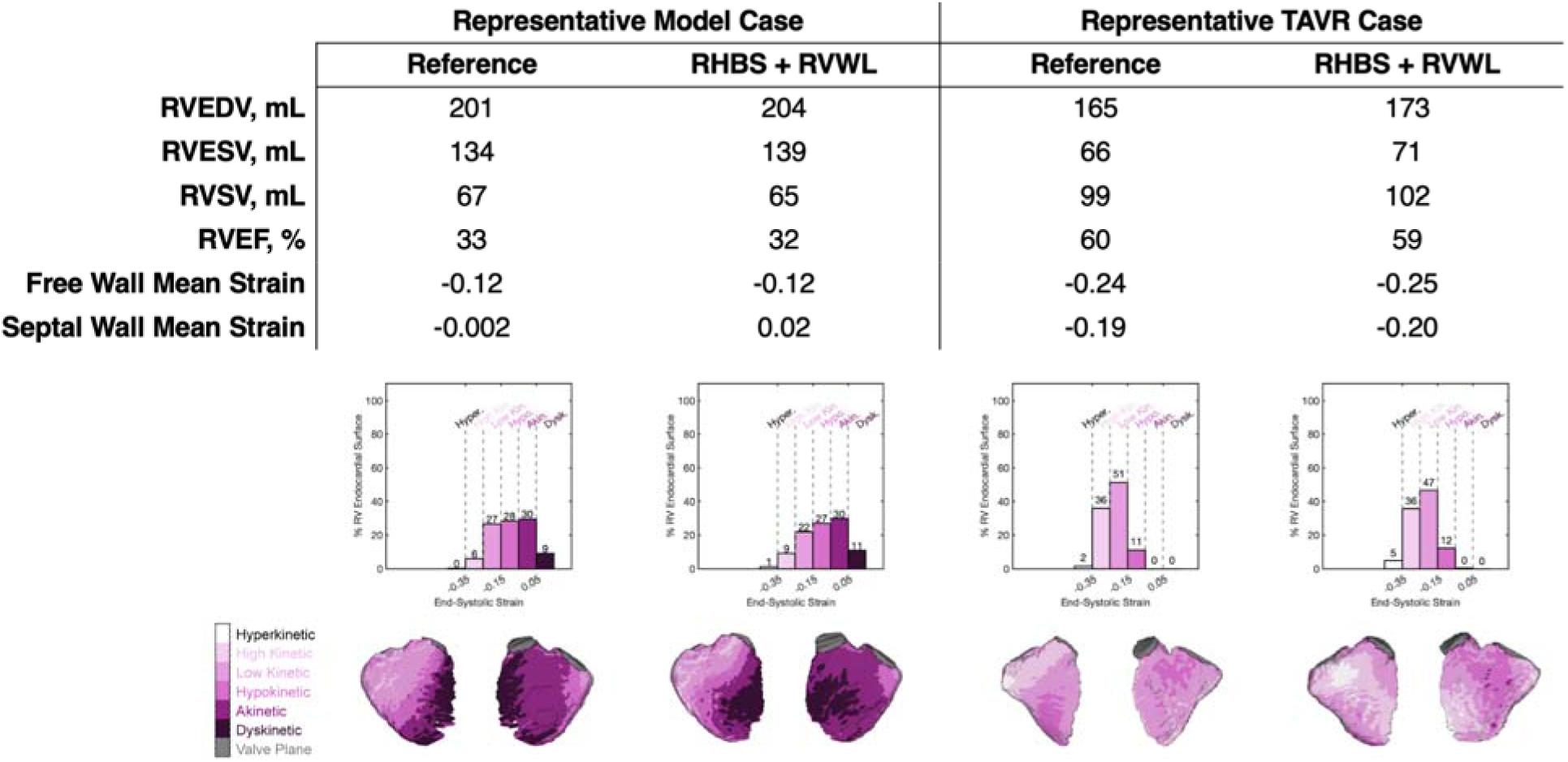
Accuracy of Quantitative RV Analysis with RHBS+RVWL-derived metrics. **Left:** Results from a representative cross validation patient. Volumetry errors due to RHBS+RVWL-based analysis in this LVHF case were 3mL for RVEDV, 5mL for RVESV, 2mL for RVSV, and 1% for RVEF. Mean strain in both the free wall was the same but slightly lower in the septal wall compared to the expert annotation. The distribution of strain categories is similar to reference values (histograms and RV maps). Cosine similarity is 0.98 for the free wall and 0.99 for the septal wall. **Right:** Results from a representative independent validation (TAVR cohort) case. Volumetry errors were 8mL for RVEDV, 5mL for RVESV, 3mL for RVSV, and 1% for RVEF. Regional mean strain in the free wall and septal wall was slightly greater than the reference. The distribution of strain categories is similar to reference (histograms). Cosine similarity is 0.96 for the free wall and 0.87 for the septal wall. 3D strain maps demonstrate similar distributions.

The cross-validation case is an LVHF patient. In this patient, absolute differences between the manual and RHBS-derived segmentation was 3 mL for RVEDV, 5 mL for RVESV, 2 mL for RVSV, and 1% for RVEF. The distribution of the strain mapping histograms is similar, with both maps showing the order of the percent of the RV surface in each category from highest to lowest being Akinetic, Hypokinetic, Low Kinetic, Dyskinetic, High Kinetic, and Hypokinetic. Similarly, 3D strain mapping shows similar spatial distribution of RV strain, with large areas of Akinetic strain surrounding patches of Dyskinetic strain in the free and septal walls.

In the independent validation example, absolute differences between the manual and RHBS-derived segmentation were 8 mL for RVEDV, 5 mL for RVESV, 3 mL for RVSV, and 1% in RVEF. The distribution of RV strain matches in both maps. 3D strain maps were also similar, however the map derived from RHBS+RVWL had more hyperkinetic areas in the free wall and hypokinetic areas in the septal wall than the reference method.

## Discussion

In this work, we describe how two neural networks, RHBS and RVWL, trained across a diverse set of RV phenotypes, can be combined to automate regional RV strain assessment. RHBS enables automated RV bloodpool segmentation while RVWL labels free and septal wall points on the RV surface. After cross-validation, we analyzed an independent cohort of patients - a series of patients undergoing TAVR - to test prospective utility. In both cohorts, the combined pipeline yielded volumetry, mean RV FW and SW strains, and a distribution of strain for each wall which closely matched the standard processing pipeline. This is a significant advancement as automated analysis of the RV has been limited by the lack of high throughput methods and need for manual inputs.

Deep learning algorithms have previously proven useful in predicting 3D medical image segmentation. In CT, whole heart segmentation models (13,15) have been developed and RV accuracy has been assessed in patients being evaluated for TAVI (14). The whole heart models have yielded RV segmentation Dice scores above 0.80. While nnU-Net has the option to perform image patching to reduce the image size and subsequent memory requirements (13–15), our implementation did not include nor require patching. While this reduces the number of training examples, it enables more straightforward processing. Alternative to patching, our group has previously demonstrated that subsampling the image based on regions of similar intensity improves processing time without sacrificing image and segmentation fidelity (12). However, this requires preprocessing and modification of the conventional network and can be offset by the availability of higher RAM GPUs.

The versatility of nnU-Net has allowed for extensive deep learning approaches to medical imaging analysis for both cardiac (17,24–26) and non-cardiac (27–30) focused applications. Prior efforts to build RV segmentation models have utilized short-axis (16), long axis, and multi-view cardiac MRI images (17,24). Martín-Isla et al and Punithakumar et al similarly demonstrated that nnU-Net was effective at predicting RV segmentations, even in cases with different pathologies (17,24), acquired at several centers and with different scanners (17). They demonstrated that segmentations predicted at the end-systolic phase were more prone to error than end-diastolic segmentations (17). We also observed success across varied pathology and observed lower Dice scores for end-systolic frames compared to end-diastolic frames (**Figure 3**). Lastly, our paper provides further evidence of the nnU-Net’s utility as prior studies have not directly reported on the ability of DL-derived segmentations to enable accurate assessment of RV function (limiting their analysis to Dice metrics.

Previously, PointNet-based architectures have been applied to cardiac imaging problems. Specifically, they’ve been used to improve 3D reconstruction of the ventricular surface on MRI (31–33) predict cardiac deformations observed in large MRI databases (34), and delineate complex structures such as the LAA in detail from CT images (35). In our work, we leverage the labeling capabilities to automate delineation of free and septal wall regions of the RV. Further work could aim to subdivide the walls into AHA segment size regions but this would require a method to reliably label subregions manually (for training purposes). Another alternative would be to define rules to subdivide the labeled areas. For example, the region labeled the free wall could be divided into basal, mid, and apical regions based on its extent along the long axis.

In this work, we pair segmentation and labeling with assessment of volumetry and regional endocardial strain assessment. CT-based endocardial strain has been shown to be reproducible (36), comparable to MR strain (20,37), and robust to low dose imaging (21,37). CT-based assessment of RV endocardial strain has demonstrated that regional RV strain from CT improves pre-operative risk scoring in complex patients (10), has been shown to match clinical profiles of RV dysfunction in several pathophysiologies (9), and can be combined with hemodynamics to estimate myocardial work (9). However, as described earlier, earlier methodology relies on time consuming, expert CT image segmentations. While our group has established automated LV segmentation models (11), RV assessment has relied on semi-automated methods. Here, we’ve demonstrated that a DL-approach, RHBS, can automate RV segmentation, and that the RV strain profiles generated from predicted segmentations are comparable to strain profiles from manual RV segmentation. RHBS will allow for functional assessment in larger populations since throughput can be significantly increased. This creates the opportunity for larger, multi-center studies with high reproducibility.

Despite these findings, this study has several limitations. First, this was a single center study where all scans were performed on a single CT scanner and all manual segmentations used for training were generated by one trained user. Given the lack of standardized training for RV bloodpool segmentation from cineCT, the time required to train others to an appropriate level of expertise would have limited the progress of this study. Second, while the training cohort included patients with a range of complex anatomies, the study lacks a true “normal” dataset. The scans included in this study were retrospectively acquired and patients with normal cardiac function are rarely indicated for cardiac CT. Analysis of the automated pipeline in comparison to other imaging modalities such as CMR and echo is left for future work. Third, while we validated our findings in an independent cohort, the patients had a separate etiology from the training cohort. They were being evaluated for planning of TAVR due to aortic stenosis. While this reflects a clinical cohort of interest where RV function has been described as an important factor, it does not directly match the variety of anatomy included in training. Therefore, future work is needed to test RHBS and RVWL’s ability in other cohorts. Fourth, while training cases were scanned at the clinical range of enhancement, we selected cases from our database of TAVR studies to include a wide range of RV enhancements. This enabled us to evaluate whether RHBS and RVWL were robust to imaging with different enhancement values (**Figure 3**). Lastly, given the success of our RHBS and RVWL models, there is an opportunity to prospectively predict RV segmentation in new populations and scale our studies to include much larger cohorts (n = 100s). However, the ability of this approach to be translated to analyze other chambers (e.g., left atrium) is unknown.

## Conclusion

In this work, we present a fully automated pipeline to perform volumetry and regional strain analysis of the RV from contrast enhanced, ECG-gated cineCT images. We developed and cross-validated two DL-based networks (RHBS and RVWL) in a diverse population of RV pathologies and then demonstrated clinical utility in an independent testing cohort.

## Data Availability

Research code and deidentified patient data reported in this article will be made available upon acceptance for publication in a Github repository.

## Acknowledgements

We’d like to thank Maria J. Ledesma-Carbayo for her insights and discussion of segmentation approaches.

## References

1. Otto CM, Nishimura RA, Bonow RO, Carabello BA, Erwin JP, Gentile F, et al. 2020 ACC/AHA Guideline for the Management of Patients With Valvular Heart Disease: A Report of the American College of Cardiology/American Heart Association Joint Committee on Clinical Practice Guidelines. Circulation [Internet]. 2021 Feb 2 [cited 2024 Oct 29];143(5). Available from: https://www.ahajournals.org/doi/10.1161/CIR.0000000000000923

2. Stout KK, Daniels CJ, Aboulhosn JA, Bozkurt B, Broberg CS, Colman JM, et al. 2018 AHA/ACC Guideline for the Management of Adults With Congenital Heart Disease: A Report of the American College of Cardiology/American Heart Association Task Force on Clinical Practice Guidelines. Circulation [Internet]. 2019 Apr 2 [cited 2021 Sep 13];139(14). Available from: https://www.ahajournals.org/doi/10.1161/CIR.0000000000000603

3. Généreux P, Pibarot P, Redfors B, Mack MJ, Makkar RR, Jaber WA, et al. Staging classification of aortic stenosis based on the extent of cardiac damage. European Heart Journal. 2017 Dec 1;38(45):3351–8.

4. Stout KK, Daniels CJ, Aboulhosn JA, Bozkurt B, Broberg CS, Colman JM, et al. 2018 AHA/ACC Guideline for the Management of Adults With Congenital Heart Disease: A Report of the American College of Cardiology/American Heart Association Task Force on Clinical Practice Guidelines. Circulation [Internet]. 2019 Apr 2 [cited 2024 Oct 29];139(14). Available from: https://www.ahajournals.org/doi/10.1161/CIR.0000000000000603

5. Goo HW. Changes in Right Ventricular Volume, Volume Load, and Function Measured with Cardiac Computed Tomography over the Entire Time Course of Tetralogy of Fallot. Korean J Radiol. 2019;20(6):956.

6. Scott A, Kligerman S, Hernandez Hernandez D, Kim P, Tran H, Pretorius V, et al. Preoperative Computed Tomography Assessment of Risk of Right Ventricle Failure After Left Ventricular Assist Device Placement. ASAIO Journal. 2023 Jan;69(1):69–75.

7. Scott A, Chen Z, Hernandez Hernandez D, Kligerman S, Kim P, Tran H, et al. Pressure Volume Loop Analysis of the Right Ventricle in Heart Failure With Computed Tomography. ASAIO Journal. 2023 Feb;69(2):e66–72.

8. Contijoch FJ, Groves DW, Chen Z, Chen MY, McVeigh ER. A novel method for evaluating regional RV function in the adult congenital heart with low-dose CT and SQUEEZ processing. International Journal of Cardiology. 2017 Dec;249:461–6.

9. Craine A, Scott A, Desai D, Kligerman S, Adler E, Kim NH, et al. Threeldimensional regional evaluation of right ventricular myocardial work from cine computed tomography: A pilot study. Medical Physics. 2025 Jun;52(6):4205–21.

10. Scott A, Chen Z, Kligerman S, Kim P, Tran H, Adler E, et al. Regional Strain of Right Ventricle From Computed Tomography Improves Risk Stratification of Right Ventricle Failure. ASAIO Journal. 2024 May;70(2):358–64.

11. Chen Z, Rigolli M, Vigneault DM, Kligerman S, Hahn L, Narezkina A, et al. Automated cardiac volume assessment and cardiac long-and short-axis imaging plane prediction from electrocardiogram-gated computed tomography volumes enabled by deep learning. European Heart Journal - Digital Health. 2021 Jun 29;2(2):311–22.

12. Gupta K, Sekhar N, Vigneault DM, Scott AR, Colvert B, Craine A, et al. Octree Representation Improves Data Fidelity of Cardiac CT Images and Convolutional Neural Network Semantic Segmentation of Left Atrial and Ventricular Chambers. Radiology: Artificial Intelligence. 2021 Nov 1;3(6):e210036.

13. Bruns S, Wolterink JM, Van Den Boogert TPW, Runge JH, Bouma BJ, Henriques JP, et al. Deep learning-based whole-heart segmentation in 4D contrast-enhanced cardiac CT. Computers in Biology and Medicine. 2022 Mar;142:105191.

14. Sharobeem S, Le Breton H, Lalys F, Lederlin M, Lagorce C, Bedossa M, et al. Validation of a Whole Heart Segmentation from Computed Tomography Imaging Using a Deep-Learning Approach. J of Cardiovasc Trans Res. 2022 Apr;15(2):427–37.

15. Dormer JD, Fei B, Halicek M, Ma L, Reilly CM, Schreibmann E. Heart chamber segmentation from CT using convolutional neural networks. In: Gimi B, Krol A, editors. Medical Imaging 2018: Biomedical Applications in Molecular, Structural, and Functional Imaging [Internet]. Houston, United States: SPIE; 2018 [cited 2024 Oct 29]. p. 100. Available from: https://www.spiedigitallibrary.org/conference-proceedings-of-spie/10578/2293554/Heart-chamber-segmentation-from-CT-using-convolutional-neural-networks/10.1117/12.2293554.full

16. Luo G, An R, Wang K, Dong S, Zhang H. A Deep Learning Network for Right Ventricle Segmentation in Short:Axis MRI. In 2016 [cited 2024 Oct 29]. Available from: http://www.cinc.org/archives/2016/pdf/139-406.pdf

17. Martín-Isla C, Campello VM, Izquierdo C, Kushibar K, Sendra-Balcells C, Gkontra P, et al. Deep Learning Segmentation of the Right Ventricle in Cardiac MRI: The M&Ms Challenge. IEEE J Biomed Health Inform. 2023 Jul;27(7):3302–13.

18. Isensee F, Jaeger PF, Kohl SAA, Petersen J, Maier-Hein KH. nnU-Net: a self-configuring method for deep learning-based biomedical image segmentation. Nat Methods. 2021 Feb;18(2):203–11.

19. Isensee F, Wald T, Ulrich C, Baumgartner M, Roy S, Maier-Hein K, et al. nnU-Net Revisited: A Call for Rigorous Validation in 3D Medical Image Segmentation. In: Linguraru MG, Dou Q, Feragen A, Giannarou S, Glocker B, Lekadir K, et al., editors. Medical Image Computing and Computer Assisted Intervention – MICCAI 2024 [Internet]. Cham: Springer Nature Switzerland; 2024 [cited 2025 Sep 3]. p. 488–98. (Lecture Notes in Computer Science; vol. 15009). Available from: https://link.springer.com/10.1007/978-3-031-72114-4_47

20. Pourmorteza A, Chen MY, van der Pals J, Arai AE, McVeigh ER. Correlation of CT-based regional cardiac function (SQUEEZ) with myocardial strain calculated from tagged MRI: an experimental study. Int J Cardiovasc Imaging. 2016 May;32(5):817–23.

21. Pourmorteza A, Keller N, Chen R, Lardo A, Halperin H, Chen MY, et al. Precision of regional wall motion estimates from ultra-low-dose cardiac CT using SQUEEZ. Int J Cardiovasc Imaging. 2018 Aug;34(8):1277–86.

22. Charles RQ, Su H, Kaichun M, Guibas LJ. PointNet: Deep Learning on Point Sets for 3D Classification and Segmentation. In: 2017 IEEE Conference on Computer Vision and Pattern Recognition (CVPR) [Internet]. Honolulu, HI: IEEE; 2017 [cited 2025 Jun 10]. p. 77–85. Available from: http://ieeexplore.ieee.org/document/8099499/

23. Qi CR, Yi L, Su H, Guibas LJ. PointNet++: Deep Hierarchical Feature Learning on Point Sets in a Metric Space [Internet]. arXiv; 2017 [cited 2025 Jun 10]. Available from: http://arxiv.org/abs/1706.02413

24. Punithakumar K, Carscadden A, Noga M. Automated Segmentation of the Right Ventricle from Magnetic Resonance Imaging Using Deep Convolutional Neural Networks. In: Puyol Antón E, Pop M, Martín-Isla C, Sermesant M, Suinesiaputra A, Camara O, et al., editors. Statistical Atlases and Computational Models of the Heart Multi-Disease, Multi-View, and Multi-Center Right Ventricular Segmentation in Cardiac MRI Challenge [Internet]. Cham: Springer International Publishing; 2022 [cited 2025 Sep 3]. p. 344–51. (Lecture Notes in Computer Science; vol. 13131). Available from: https://link.springer.com/10.1007/978-3-030-93722-5_37

25. Salimi Y, Mansouri Z, Nkoulou R, Mainta I, Zaidi H. Deep Learning-Based CT-Less Cardiac Segmentation of PET Images: A Robust Methodology for Multi-Tracer Nuclear Cardiovascular Imaging. J Digit Imaging Inform med [Internet]. 2025 May 6 [cited 2025 Sep 3]; Available from: https://link.springer.com/10.1007/s10278-025-01528-0

26. Summerfield N, Morris E, Banerjee S, He Q, Ghanem AI, Zhu S, et al. Enhancing Precision in Cardiac Segmentation for Magnetic Resonance-Guided Radiation Therapy Through Deep Learning. International Journal of Radiation Oncology*Biology*Physics. 2024 Nov;120(3):904–14.

27. Montalvo-Garcıa D, Ortuno JE, Ramos-Guerra AD, Granados-Aparici S, Goswami SS, Diaz PS, et al. Stromal Tissue Segmentation in Multi-Stained Serial Histopathological Sections of Pancreatic Tumors.

28. Ferrante M, Rinaldi L, Botta F, Hu X, Dolp A, Minotti M, et al. Application of nnU-Net for Automatic Segmentation of Lung Lesions on CT Images and Its Implication for Radiomic Models. JCM. 2022 Dec 9;11(24):7334.

29. Salimi Y, Mansouri Z, Sun C, Sanaat A, Yazdanpanah M, Shooli H, et al. Deep learning-based segmentation of ultra-low-dose CT images using an optimized nnU-Net model. Radiol med. 2025 Mar 18;130(5):723–39.

30. Pettit RW, Marlatt BB, Corr SJ, Havelka J, Rana A. nnU-Net Deep Learning Method for Segmenting Parenchyma and Determining Liver Volume From Computed Tomography Images. Annals of Surgery Open. 2022 Jun;3(2):e155.

31. Beetz M, Yang Y, Banerjee A, Li L, Grau V. 3D Shape-Based Myocardial Infarction Prediction Using Point Cloud Classification Networks. In: 2023 45th Annual International Conference of the IEEE Engineering in Medicine & Biology Society (EMBC) [Internet]. Sydney, Australia: IEEE; 2023 [cited 2025 Jul 10]. Available from: https://ieeexplore.ieee.org/document/10340878/

32. Beetz M, Banerjee A, Grau V. Point2Mesh-Net: Combining Point Cloud and Mesh-Based Deep Learning for Cardiac Shape Reconstruction. In: Lecture Notes in Computer Science [Internet]. Cham: Springer Nature Switzerland; 2022 [cited 2025 Jul 10]. p. 280–90. Available from: https://link.springer.com/10.1007/978-3-031-23443-9_26

33. Beetz M, Banerjee A, Ossenberg-Engels J, Grau V. Multi-class point cloud completion networks for 3D cardiac anatomy reconstruction from cine magnetic resonance images. Medical Image Analysis. 2023 Dec;90:102975.

34. Beetz M, Ossenberg-Engels J, Banerjee A, Grau V. Predicting 3D Cardiac Deformations with Point Cloud Autoencoders. In: Lecture Notes in Computer Science [Internet]. Cham: Springer International Publishing; 2022 [cited 2025 Jul 10]. p. 219–28. Available from: https://link.springer.com/10.1007/978-3-030-93722-5_24

35. Gao Q, Lin H, Qian J, Liu X, Cai S, Li H, et al. A deep learning model for efficient end-to-end stratification of thrombotic risk in left atrial appendage. Engineering Applications of Artificial Intelligence. 2023 Nov;126:107187.

36. Colvert GM, Manohar A, Contijoch FJ, Yang J, Glynn J, Blanke P, et al. Novel 4DCT Method to Measure Regional Left Ventricular Endocardial Shortening Before and After Transcatheter Mitral Valve Implantation. Structural Heart. 2021 Jul;5(4):410–9.

37. Manohar A, Colvert GM, Ortuño JE, Chen Z, Yang J, Colvert BT, et al. Regional left ventricular endocardial strains estimated from lowldose 4DCT: Comparison with cardiac magnetic resonance feature tracking. Medical Physics. 2022 Sep;49(9):5841–54.

